# How to measure the controllability of an infectious disease?

**DOI:** 10.1101/2023.10.10.23296471

**Authors:** Kris V Parag

## Abstract

Quantifying how difficult it is to control an emerging infectious disease is crucial to public health decision-making, providing valuable evidence on if targeted interventions e.g., quarantine and isolation, can contain spread or when population-wide controls e.g., lockdowns, are warranted. The disease reproduction number, *R*, or growth rate, *r*, are universally assumed to measure controllability because *R*=1 and *r*=0 define when infections stop growing and hence the state of critical stability. Outbreaks with larger *R* or *r* are therefore interpreted as less controllable and requiring more stringent interventions. We prove this common interpretation is impractical and incomplete. We identify a positive feedback loop among infections intrinsically underlying disease transmission and evaluate controllability from how interventions disrupt this loop. The epidemic gain and delay margins, which respectively define how much we can scale infections (this scaling is known as gain) or delay interventions on this loop before stability is lost, provide rigorous measures of controllability. Outbreaks with smaller margins necessitate more control effort. Using these margins, we quantify how presymptomatic spread, surveillance limitations, variant dynamics and superspreading shape controllability and demonstrate that *R* and *r* only measure controllability when interventions do not alter timings between the infections and are implemented without delay. Our margins are easily computed, interpreted and reflect complex relationships among interventions, their implementation and epidemiological dynamics.

## Introduction

Understanding and quantifying the effort required to control or contain outbreaks is a principal goal of infectious disease epidemiology [1]. During emergent stages of a potential epidemic, when populations are immunologically naïve, assessments of disease controllability provide critical evidence on whether targeted interventions, for example contact tracing, isolation and quarantines, are sufficient to curb spread [2] or whether non-selective control actions, such as population-level lockdowns and closures, are necessary [3]. These assessments typically rely on mathematical models [4] that combine disease surveillance data (e.g., infection times and cases) with intervention mechanisms (e.g., how isolation interrupts transmission chains), to estimate controllability (in some sense) and have informed the public health responses for influenza, measles, SARS, Ebola virus disease and COVID-19, among others [2,3,5–7].

Despite these applications, a systematic and rigorous definition of controllability is lacking [8–10]. While key factors influencing the difficulty of controlling epidemics such as transmissibility, superspreading levels, the efficiency of contact tracing and the proportion of presymptomatic infections are known [1,8,11], studies generally compute the reproduction number *R*, or (less commonly) the epidemic growth rate *r*, under proposed interventions to measure controllability [12]. For example, the impact of contact tracing and presymptomatic spread on controllability are assessed by how they effectively change *R* [1,13,14]. Here we use *R* and *r* to generally indicate the (constant) controlled reproduction number and growth rate subject to some control action or intervention. If no controls are applied, these become the popular basic reproduction number and intrinsic growth rate. The relationship between *R* and *r* depends on the pathogen generation time distribution [15], *w*, which describes the times between infections.

As *R*=1 or *r*=0 defines critical epidemic stability i.e., the state where infections will neither grow nor wane [4], it seems reasonable to base controllability on the distance of *R*-1 or *r*-0. Stable epidemics have waning infections (*R*<1, *r*<0) and unstable ones (*R*>1, *r*>0) feature exponential growth. We therefore expect that larger *R* or *r* values signify reduced controllability, justifying stronger interventions, while smaller values imply augmented robustness to transmissibility changes or intervention relaxations. The common interpretation of these distances is that we must scale infections by 1/*R* within timeframes proportional to 1/*r* to reach critical stability [12]. This interpretation further underlies related measures of intervention efficacy such as the herd immunity threshold (i.e., the proportion of the susceptible population that must be vaccinated or acquire immunity) [4,12] and the proportion of infections that must be targeted by contact tracing [2] (both relate to 1-1/*R*), as well as the speed at which isolation or digital tracing [9,13,16] must be applied to suppress infections (both relate to doubling time log(2)/*r*).

In this study we prove that the above interpretations are only valid under impractical and quite restrictive assumptions. We start by recognising that, intrinsically, an epidemic represents a positive feedback loop between past and upcoming infections (see **Fig *1*A**). Interventions are then control actions that disrupt this loop. This reframing of the disease transmission process allows us to adapt tools from control theory [17] and derive what we term as the epidemic transfer function. This captures how incident (new) infections are generated under arbitrary generation time distributions and (linear) control actions in response to imported infection time series. We propose a rigorous controllability measure defined by the gain and delay margins of the epidemic transfer function, which quantify two important and distinct distances from the critical stability point (see **Fig *1*B**). If an intervention is applied to two outbreaks, for example, the one with the larger pair of margins is more controllable under that intervention.

The gain margin *M*_*G*_ is the factor by which we can scale infections (known as the gain) before critical stability is attained, with *M*_*G*_ =1 demarcating critical stability [18]. An *M*_*G*_=2 means the epidemic remains controlled unless infections double (e.g., from releasing interventions or the emergence of more pathogenic variants), while *M*_*G*_=1/2 means we must halve infections (e.g., via more stringent interventions that reduce contact rates) to control transmission. The delay margin *M*_*D*_ quantifies the lag we can afford when imposing interventions, with *M*_*D*_ =0 delimiting critical stability [18]. An *M*_*D*_ =7 indicates that if we take more than a week to intervene (e.g., to trace and isolate infected individuals), then we are unable to keep the epidemic controlled. This (*M*_*G*_, *M*_*D*_) pair framework yields a number of advantages and controllability results.

First, our margins more accurately describe what *R* and *r* only attempt to quantify – the scale and speed of required control effort. Particularly, we show that the universal 1/*R* interpretation of controllability is only valid if interventions reduce infections without inducing dynamics and are implemented without delay. Under those conditions *M*_*G*_=1/*R* and *M*_*D*_ is unimportant (i.e., it is undefined if *R*>1 or infinite if *R*<1). However, this is unrealistic given mounting evidence that interventions change generation time and other epidemiological distributions (which is how a control action induces dynamics) and that practical constraints on outbreak control inevitably cause lags [7,19–22]. Additionally, by interrogating our epidemic transfer function, we find that *r* only quantifies the asymptotic epidemic growth rate [23] and so neglects short-term dynamics (which are crucial for understanding unwanted oscillations in infections) and their interactions with imposed interventions. These effects belie our conventional notions of controllability.

Second, our margin-based framework generalises and unifies earlier approaches [1,8]. We characterise how presymptomatic spread, transmission heterogeneities from superspreading, multitype epidemics (but without considering contact structures) or co-circulating variants and surveillance limitations (e.g., reporting delays and underreporting) all modulate controllability. These complexities can be commonly evaluated using our two margins, which always have the same interpretation. This is beneficial because *R* or *r* is not always clearly defined or even meaningful for some of these complexities [24,25]. Importantly, our margins yield thresholds of controllability under these complexities that can be directly compared to decide the relative effectiveness of targeted and population-level interventions. These thresholds reduce to more conventional 1-1/*R* type results under the restrictive conditions mentioned above.

Last, our margins offer a more complete measure of controllability. Because induced dynamics from interventions, implementation delays and surveillance imperfections are pervasive, even if proposed interventions are expected to drive *R*<1 or *r*<0, this does not reliably inform about the required control effort and the robustness of the epidemic once controlled by these actions. We find ample evidence of controlled (*R*<1) epidemics with *M*_*G*_<1/*R*, indicating that standard controllability interpretations overestimate robustness to increases in infections. We also show that some of these controlled epidemics possess *M*_*D*_=7-14 days, signifying that if the combined lag from surveillance and intervention delays rises above this value (e.g., due to downscaling of surveillance or control programmes) then the epidemic will become destabilised. Neither *r* nor *R* can generally expose these issues. Our methodology probes the notion of controllability and raises questions about the understudied rebound effects of interventions.

## Methods

### Renewal models and transfer functions

The renewal branching process [26] is a fundamental and popular infectious disease model that has been applied to describe epidemics of COVID-19, pandemic influenza, Ebola virus disease, measles, SARS and many others [12,27]. This model defines how incident infections at time *t, i*(*t*) depend on the reproduction number *R* and incidence at earlier times *i*(*τ*) (*τ* ≤ *t*, with a limit infinitesimally before *t*) via the autoregressive relationship in the left of **Eq. (M1)**.

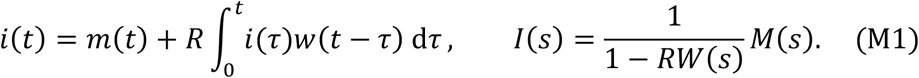

We assume that *R* is constant during our period of interest. As **Eq. (M1)** has no interventions and we consider initial epidemic stages, *R* is the basic reproduction number. Note that *R* can describe other (constant) effective reproduction numbers when the renewal process is a good approximation to later epidemic stage dynamics. Because this model is linear, we neglect non-linear effects such as those due to the depletion of individuals that are susceptible to infection.

**Eq. (M1)** includes input infections *m*(*t*) that have been imported or introduced into our region of interest and which eventually contribute to onwards transmission [28]. Our output is *i*(*t*). The kernel of the renewal autoregression is *w*(*t* − *τ*), which is the probability of an infection being transmitted after a duration of *t* − *τ* time units. The set of coefficients {*w*(*k*), *k* ≥ 0} composes the generation time distribution of the disease [15]. This captures variability in the time it takes for a primary infection to cause a secondary one. The generation time distribution is a key characteristic of a pathogen that determines the temporal aspects of its spread via the convolution in **Eq. (M1)**. We denote the mean generation time as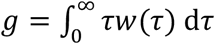.

Since **Eq. (M1)** is a linear model, we can analyse it in the frequency or *s* domain using Laplace transforms e.g., 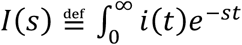 d*t* is the transform of *i*(*t*). This gives the right side of **Eq. (M1)** after some algebra with capitalised forms as the transformed version of variables from the time domain. We visualise this using the block diagram of **Fig 1A**. We propose the ratio *G*(*s*) = *I*(*s*)*M*(*s*)^−1^ as the epidemic transfer function (TF) that maps input importations onto output infections. The roots of its characteristic polynomial 1 − *RW*(*s*) (the denominator of *G*(*s*)) are the poles of the renewal process and completely define the stability of the epidemic [17]. A stable epidemic (infections decay with time given initial imports) has poles with negative real parts. An unstable epidemic (infections grow) has at least one pole with positive real part. Critical stability requires at least one pole with real part of 0, with all others negative.

**Fig 1:**
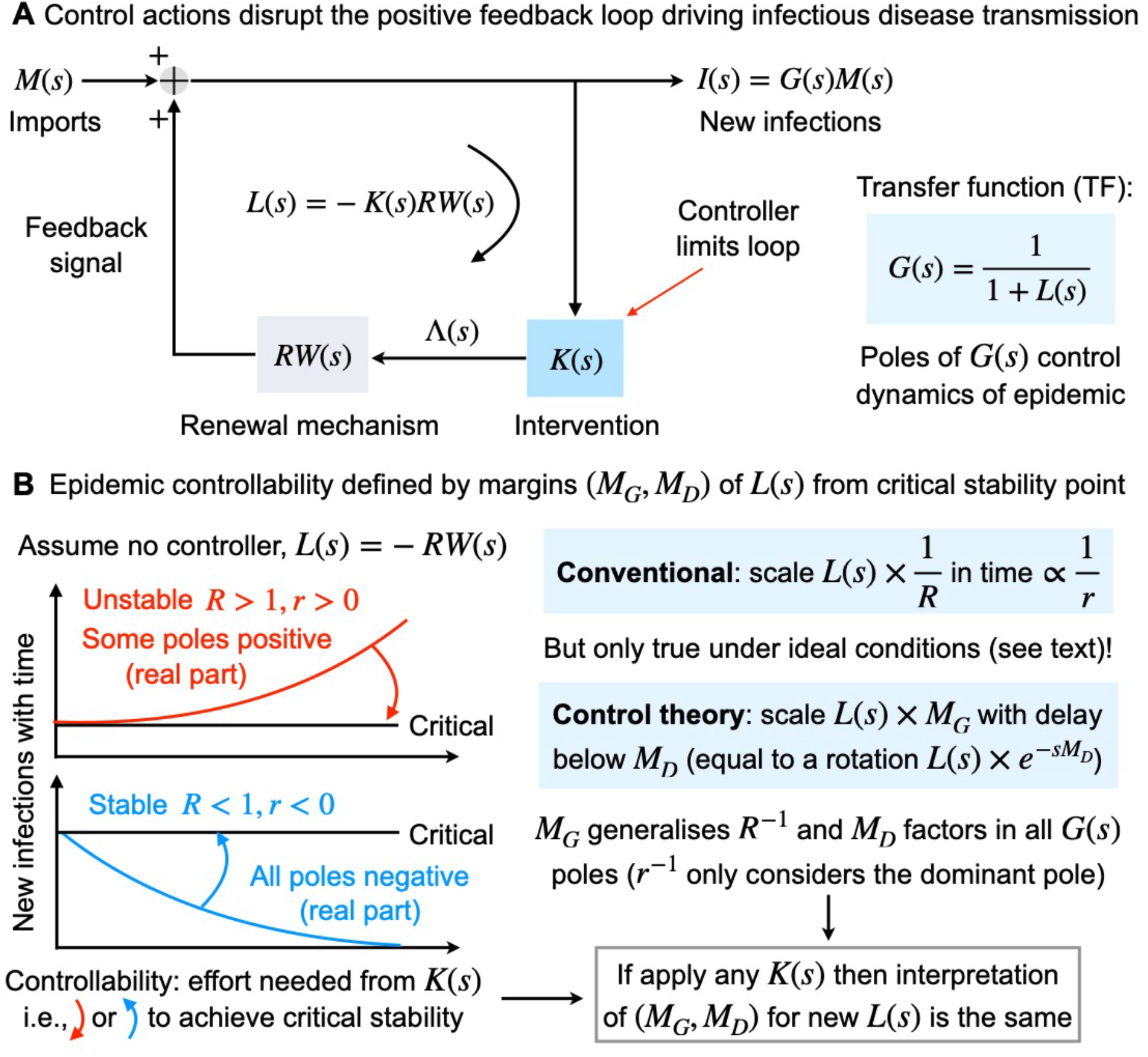
Epidemic architecture and controllability definition. Panel A shows that the renewal epidemic model is a positive feedback system with signals being successively fed back along a loop and added to imported infections *M*(*s*). The loop TF *L*(*s*) (negative by convention for positive feedback) governs the poles of the closed loop TF *G*(*s*), which completely expresses how imports combine with the epidemic dynamics to generate new infections *I*(*s*). When we intervene or initiate control action, we disrupt the feedback loop via a controller *K*(*s*). Panel B explains the concept of controllability. This is the effort needed to drive the epidemic to critical stability where at least one pole has a 0 real part (others must be negative). We sketch stable and unstable epidemics with no controller and an initial import and contrast the conventional notion of controllability with our control theoretic approach. Our margin pair completely and precisely describe how *L*(*s*) can be forced to criticality in the complex plane (via scaling and rotation) and, crucially, emphasise that the distance of *L*(*s*) from −1 measures controllability. The distances of *r* from 0 and *R* from 1 are restricted specialisations of this condition,

The form of the characteristic polynomial of *G*(*s*) confirms that the dynamics of the epidemic depend explicitly on *R* and the generation time distribution. These are two of three quantities commonly used to depict the transmissibility of infectious diseases. The third is the asymptotic exponential growth rate of infections, 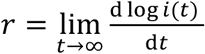 and also emerges from **Eq. (M1)**. Since *W*(−*s*) is equivalent to the moment generating function of the generation time distribution evaluated at *s*, we know from [15] that *W*(*r*) = *R*^−1^. Interestingly, *r* is also the dominant pole of *G*(*s*). Often the growth rate is expressed as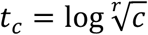, the time it takes for infections to (asymptotically) grow (or decline) by a factor *c*. At *c* = 2 we get the popular epidemic doubling time. We compute appropriate forms of *G*(*s*) and its poles for generalisations of **Eq. (M1)** that model various interventions under practical constraints in the Results.

### Generation time distributions and Laplace transforms

The dynamics of infectious diseases are largely determined by the generation time distribution because *W*(*s*) is the only non-constant term in the TF of **Eq. (M1)**. We model *W*(*s*) as a phase-type distribution, which is an expansive class built from combinations and convolutions of exponential distributions. This class can approximate any distribution [29] and includes the Erlang, exponential, deterministic (degenerate) and bimodal distributions that we examine in the Results. Erlang (or related gamma), deterministic and exponential distributions are used to model influenza, measles and COVID-19, among others [3,15,26,27]. Multimodal and mixture distributions are applied to diseases featuring multiple stages (which may even involve vectors) or pathways of transmission, such as malaria and Ebola virus disease [19,30].

All phase-type distributions conform to the relations in **Eq. (M2)**, where we use bold to denote vectors or matrices and ***x***^1^ to denote the transpose of some row vector ***x***.

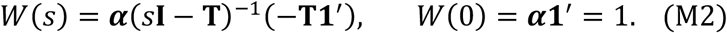

In **Eq. (M2) T** is a *n*^2^ matrix of transition rates among the *n* distribution states, **I** the *n*^2^ identity matrix, ***α*** is a row vector of length *n* summing to 1 (providing weights to the states) and **1 is a ro**w vector of *n* ones. Here *n* represents the complexity of the phase type distribution relative to how exponential distributions are combined. A standard exponential distribution has *n* = 1. Mixtures of phase-type distributions are also phase-type and we observe that their Laplace transforms evaluate to 1 at *s* = 0 (equivalent to the fact that probability distributions integrate to 1). We find that this basic property important for computing controllability later in the Results.

For a mean generation time *g* we can construct an Erlang distribution with shape *a* and scale *b* such that *g* = *ab* by setting *n* = *a*, ***α*** = [1 0 … 0], and **T** as a matrix with non-zero elements of **T**_*kk*_ = −*b*^−1^ and **T**_*kk*+1_ = *b*^−1^ for 1 ≤ *κ* ≤ *n*. As a result, we obtain *W*(*s*) as in **Eq. (M3)**.

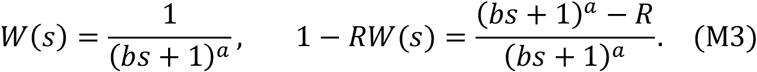

We also find the characteristic polynomial or denominator from **Eq. (M1)**. This has roots when 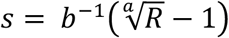, which is the formula for the growth rate as expected from [15]. Exponential and deterministic distributions have *a* = 1 and *a* → ∞, respectively. We get the roots of the characteristic polynomial of the exponential distribution by simply substituting in **Eq. (M3)**. The deterministic distribution yields *W*(*s*) = *e*^−*sg*^ at the limit, is equivalent to applying a delay of *g* time units and has solution to its characteristic polynomial of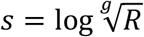.

The bimodal distribution we consider is a mixture of two Erlang distributions with state sizes *n*_1_ = *a*_1_ and *n*_2_ = *a*_2_ and ***α*** = [*α*_1_ 0 … 0, 1 – *α*_1_ 0 … 0], which has *n*_1_ − 1 and then *n*_2_ – 1 zeros respectively. The choice of *α*_1_ defines the mixture weighting. The state matrix has size (*n*_1_ + *n*_2_)^2^ with 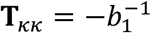 and 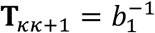 for 1 ≤ *k* ≤ *n*_1_ and 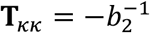 and 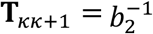 for *n*_1_ + 1 ≤ *k* ≤ *n*_2_. The *b*_1_ and *b*_2_ are chosen to get mean generation time *g*. We obtain 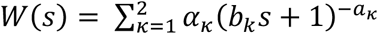 and numerically compute roots of its characteristic polynomial.

We can easily extend this formulation to allow higher order mixtures. The phase-type structure allows us to describe complex distributions without losing analytical tractability.

### Margins of stability and notions of controllability

In the above subsections we described the elements of the renewal epidemic model and its characteristic polynomial 1 − *RW*(*s*). Here we review the concepts of gain, phase and delay margin from classical control theory, which underpin our Results and provide measures of how distant linear systems are from critical stability [17]. The loop TF *L*(*s*) = −*RW*(*s*) (under no control) captures the dynamics around the loop as in the block diagram of **Fig 1A**. While in the Results we expand this *L*(*s*) formulation to include a controller *K*(*s*) that describes our epidemic intervention and investigate more generalised model architectures, the principles and interpretation that we detail here remain valid for all of these complexities.

Using *L*(*s*) our characteristic polynomial becomes 1 + *L*(*s*) with the epidemic TF as *G*(*s*) = X1 + *L*(*s*)Y^−1^. Poles are complex solutions to *L*(*s*) = −1 + *j*0 where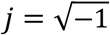. We can write a pole as the complex number *σ* + *jω*. At the critical stability point the dominant pole has *σ* = 0 so that *L*(*jω*) = −1. Control theory [17] states that the distance in the complex plane of *L*(*jω*) from −1 reflects the stability properties of the process. We can describe this distance by the multiplicative factor (the gain), and the angular change (the phase) that respectively scale and rotate *L*(*jω*) onto −1 + *j*0 in the complex plane. These distances are known as the gain and phase margin [17] and relate to polar descriptions of complex numbers. Note that the other poles also contribute to the form of *L*(*jω*) and so influence the margins.

The gain margin 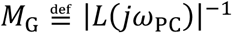 is the inverse of the magnitude of *L*(*s*) evaluated at *ω*_PC_ the first frequency at which the phase crosses −*π* radians. Here |. | denotes magnitude so 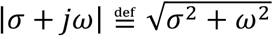. The phase margin 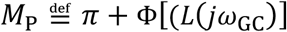 is *π* plus the phase Φ[.] (in radians) evaluated at *ω*_GC_, the frequency where |*L*(*jω*)| first crosses 1 from above (known as gain crossover). This measures how much phase lag (i.e., clockwise rotation in the complex plane) can be added to *L*(*jω*_GC_) before driving the epidemic to critical stability [31,32]. Phase margin is not intuitive for our analyses but can be transformed into a more interpretable delay margin *M*_D_ (e.g., in some cases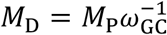[31]). This margin quantifies how much pure time delay or lag forces *L*(*s*) to the critical point (lag reduces phase).

We compute both margins using in-built functions (specifically allmargin(.)) from the MATLAB control system toolbox (see Data Availability). This essentially evaluates the magnitude and phase of *L*(*s*) at every *s* = *jω* and finds the appropriate crossover frequencies for determining the margins. When systems have multiple *ω*_PC_ or *ω*_GC_ crossover frequencies we consider the minimum margin to ensure our characterisation is robust. The gain and delay margins define the two ways (scaling and rotation) that we can drive our epidemic to critical stability. If we multiply *L*(*s*) by the scale factor *M*_G_ or the angular shift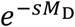, which is the Laplace transform of a pure delay of *M*_D_ time units, we drive the epidemic to the critical stability point [32]. We compare this interpretation to more conventional epidemic measures in **Fig 1B**.

Importantly, the distance of our system from stability requires specifying both the gain margin and the delay margin [17]. We propose this pair representation as our measure of epidemic controllability, which quantifies the intervention effort required to control an unstable epidemic or the perturbation (e.g., to disease transmissibility *R*) required to destabilise an epidemic that is already under control. We expand this definition to include various model architectures and interventions in the Results. Note that in control theory, a system is only formally controllable if inputs exist that drive it from any initial state to any desired state in finite time. Our definition is more relaxed and only considers what changes force epidemics to critical stability and the required intensity of those changes. This relates to margins (also termed relative stability) and aligns with the informal definition commonly applied in infectious diseases [1,10].

Although controllability here defines the control effort needed to stabilise infections, it does not measure performance. Performance depends on our control objectives i.e., what we want our interventions to achieve [17]. These may include desired margins but generally we may want our system response, *i*(*t*) in this setting, to meet some desired dynamics *u*(*t*). A key long-term performance metric is the error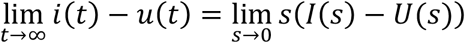. The second limit follows from the final value theorem [17]. If *U*(*s*) = *us*^−1^ is a desired equilibrium of infections (*us*^−1^ is the Laplace transform of a step function with amplitude *u*), then 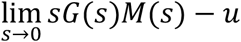 measures accuracy. Important short-term performance measures are the peak overshoot 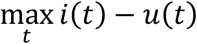 and the level of oscillation of *i*(*t*) about either *u*(*t*) or the equilibrium.

## Results

We start by expanding the framework in the Methods to explore the standard assumption that larger *R* or *r* signals a less controllable epidemic [1,10,33]. This belief is sensible as increases in *R* cause infections to multiply more and rises in *r* engender faster multiplication. In **Fig 1B** we recap these conventional notions of controllability. We prove that these universal notions are only true under restrictive and idealistic intervention assumptions. Extending the margins above, we construct a rigorous measure of epidemic controllability that accurately reflects the control effort needed to stabilise a growing epidemic and the robustness to perturbations of a controlled epidemic. To maintain analytic tractability and as we focus on deriving fundamental insight into controllability, we study constant *R* or *r* and neglect stochasticity (see Methods).

In later sections we discuss relaxations of these assumptions and show that our framework can assess how variant dynamics, presymptomatic spread, superspreading, intervention lags and surveillance biases all modulate controllability.

### Epidemic models, feedback control and transfer functions

The renewal process is widely used to model acute infectious diseases and is given in **Eq. (M1)** of the Methods. There new or output infections at time *t, i*(*t*) result from multiplying all active infections by *R* with *m*(*t*) as the introduced infections or input. Past infections are active if they can still transmit. The generation time distribution {*w*(*k*), *k* ≥ 0} sets the transmission probabilities [26,27] with the active infections computed as a convolution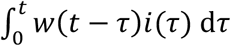.

However, these quantities from **Eq. (M1)** describe uncontrolled dynamics. Here we extend this model to include control. We define a generic control strategy as one reducing infections to *λ*(*τ*) ≤ *i*(*τ*) so that 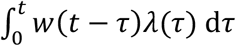 is the equivalent convolution. This yields **Eq. (1)**.

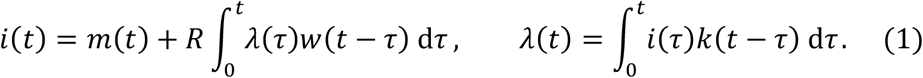

The controller achieves this reduction by weighting past infections by a kernel *k*(*τ*) with overall effect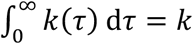. If this kernel only has mass at the present so *k*(*t*) = *kδ*(*t*), with *δ*(*t*) as the Dirac delta function, we get constant (memoryless) feedback control and *λ*(*t*) = *ki*(*t*). Generally, 0 ≤ *k* ≤ 1 as control reduces infections. However, if the epidemic is already stable, we may set *k* > 1 to assess robustness to perturbations in infections i.e., we want to find the largest *k* that achieves critical stability. The expressions in **Eq. (1)** become the standard ones of **Eq. (M1)** by removing control i.e., by using a constant controller with overall effect *k* = 1.

We Laplace transform **Eq. (1)** (see Methods) with *I*(*s*), *M*(*s*) and *W*(*s*) as the transformed infection incidence, importations and generation time distribution in the frequency or *s* domain. Since convolutions are products in this domain, the controller satisfies Λ(*s*) = *K*(*s*)*I*(*s*) with *K*(*s*) as its transform. We represent these operations as the block diagram in **Fig 1A**, where we identify that, fundamentally, an epidemic involves a positive feedback loop between past and upcoming infections. Taking the product of blocks along the loop we obtain the loop transfer function (TF) as *L*(*s*) in **Eq. (2)** below. Control aims to disrupt this loop.

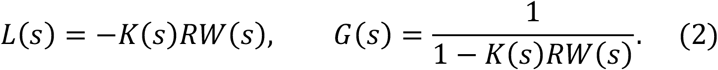

Using this structure, we can define transmission dynamics by the properties of the closed loop TF, *G*(*s*) = *I*(*s*)*M*(*s*)^−1^, which describes how imports drive total incidence. We can see the importance of *L*(*s*) by noting that *G*(*s*) = X1 + *L*(*s*)Y^−1^ [18]. The poles of *G*(*s*) determine the dynamics and stability of the epidemic and are complex number solutions of *L*(*s*) = −1 + *j*0 (see Methods). We recover the uncontrolled epidemic TFs by setting *K*(*s*) = 1, which is the Laplace transform of the constant controller above with *k* = 1.

We can interpret **Eq. (2)** by recognising that an unstable epidemic (at least one pole of *G*(*s*) has a positive real part) successively multiplies infections along the loop. This constitutes the positive feedback we illustrate in **Fig 1A**. Interventions or control actions having magnitude |*K*(*s*)| < 1 limit this positive feedback by interfering with the loop and hence attenuating this multiplication. Modification of the intrinsic epidemic dynamics *RW*(*s*) by the controller *K*(*s*) within the loop achieves this goal. A stable epidemic (all poles of *G*(*s*) have non-positive real parts) is also multiplicative, but infections reduce along the loop. We can apply |*K*(*s*)| > 1 as an amplifier of infections to study the robustness of the controlled epidemic to any destabilising perturbations or uncertainties (e.g., surges in transmissibility or more pathogenic variants).

There are two important corollaries of **Eq. (2)**. First, the poles of the epidemic TF *G*(*s*) are the roots of the characteristic polynomial 1 − *K*(*s*)*RW*(*s*). Solving this (see Methods), we find the epidemic growth rate *r* is the dominant pole i.e., it is the major contributor to the dynamics of the system (see Methods for explicit calculation in the uncontrolled case) and its variations reflect the impact of the controller *K*(*s*). Second, *K*(*s*) directly regulates both the generation times and *R*. For constant controller *K*(*s*) = *k*, the epidemic has an effective reproduction number of *kR*, with related changes to its effective growth rate. These observations seemingly support the common paradigm of modelling interventions and assessing controllability directly from how *R* or *r* (or related parameters such as doubling times or infectiousness) change [12].

### A framework for investigating epidemic controllability

However, these corollaries actually expose why these parameters are insufficient for defining controllability i.e., the effort required to stabilise an unstable epidemic, or the intensity of the perturbations required to destabilise a stable epidemic. Specifically, the difficulty of controlling the epidemic in real time also depends on its other poles (which may be oscillatory) [18] and only asymptotically are infections completely determined by the dominant pole *r*. Additionally, the assumption that *K*(*s*) is constant and introduces no dynamics is unrealistic (e.g., isolation is known to reduce generation times [20,21]) and only likely true in very limited circumstances. We therefore need to account for transient and intervention-induced dynamics [23,34].

To investigate the implications of these corollaries, we propose a new framework for defining epidemic controllability, which adapts classical control theory as well as generalises and more rigorously quantifies the interpretation frequently ascribed to *R* or *r*. **Fig 1B** summarises and contrasts this framework to standard (albeit implicit) notions of controllability. We know from **Eq. (2)** and stability theory [17] that as *L*(*s*) approaches −1 + *j*0 in the complex plane the closed loop *G*(*s*) becomes critically stable i.e., it is on the verge of instability with *r* = 0. The gain *M*_G_ and delay *M*_D_ margins [18] precisely determine the distance of *L*(*s*) from −1 + *j*0 (see Methods for how to compute these and related margins) [17].

For stable epidemics (*r* < 0 i.e., all *G*(*s*) poles are in the left half of the complex plane), *M*_*G*_ and *M*_>_ respectively measure how much we can scale up or delay infections before the system becomes critical [32]. Accordingly, for unstable epidemics (*r* > 0 i.e., at least one *G*(*s*) pole is in the right half plane) they quantify how much we must scale down or limit delay to stabilise an epidemic (assuming certain conditions [17]). If an epidemic admits a margin pair (*M*_D_, *M*_G_), then we can multiply *L*(*s*) along its loop by 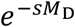 or *M*_*G*_ respectively to force the epidemic to its critical stability point. Critical stability is rigorously defined as when *K*(*s*) = 1 in **Eq. (2)** then *r* matches *R* − 1 in sign and is the dominant *G*(*s*) pole so *L*(*s*) = −1, *R* = 1 and *r* = 0 all correspond. There is an analogous association with the effective *R* and *r* when some control is acting (*K*(*s*) ≠ 1). The crucial distinction we make is that the distance of *L*(*s*) from −1 and not that of *R* from 1 or *r* from 0 is what actually determines controllability.

The margins we propose to measure this distance precisely and holistically characterise the essence of earlier notions of control effort by quantifying the magnitude and time by which we must alter infections to attain the brink of stability. Stable or controlled epidemics feature *M*_G_ > 1 and *M*_D_ > 0 and larger margins signify better controllability (see **Fig 1B**). Computing these for **Eq. (2)**, we get *M*_G_ = |−*K*(*jω*_PC_)*RW*(*jω*_PC_)|^−1^ [17] where *ω*_PC_ is the phase crossover frequency (see Methods). From the properties of distributions, *W*(0) = 1. We confirm this in the Methods for a universal class of phase-type generation time distributions [29] that include realistic models of *w*(*t*) for many infectious diseases [15,27]. Accordingly, when *ω*_PC_ = 0, *M*_G_ = |−*K*(0)*R*|^−1^. As critical stability occurs when *M*_G_ = 1, the critical control effort required, based on this gain margin, is therefore *K*^*^ = |*K*(0)| = *R*^−1^.

We show in **Fig 2** for constant controllers applied to epidemics with various generation time distribution shapes (**Fig 2A**) that *ω*_PC_ = 0 is true and unique. For stable epidemics, we find *M*_D_ → ∞ (not shown but see Data Availability for linked code). Consequently, under these conditions, controllability is completely established by the magnitude of *R*^−1^ (**Fig 2C**), which correlates well with the Euclidean distance in the complex plane between *L*(*s*) and −1 (inset). When the epidemic is unstable the gain margin is also set by *R*^−1^ but there may be ways of removing system lag that also define a dimension of control. However, if we apply a constant controller (so system lag does not change) with *k* = *αR*^−1^ and *α* < 1, the controlled epidemic has an effective reproduction number of *α* and hence a controllability of *α*^−1^.

**Fig 2:**
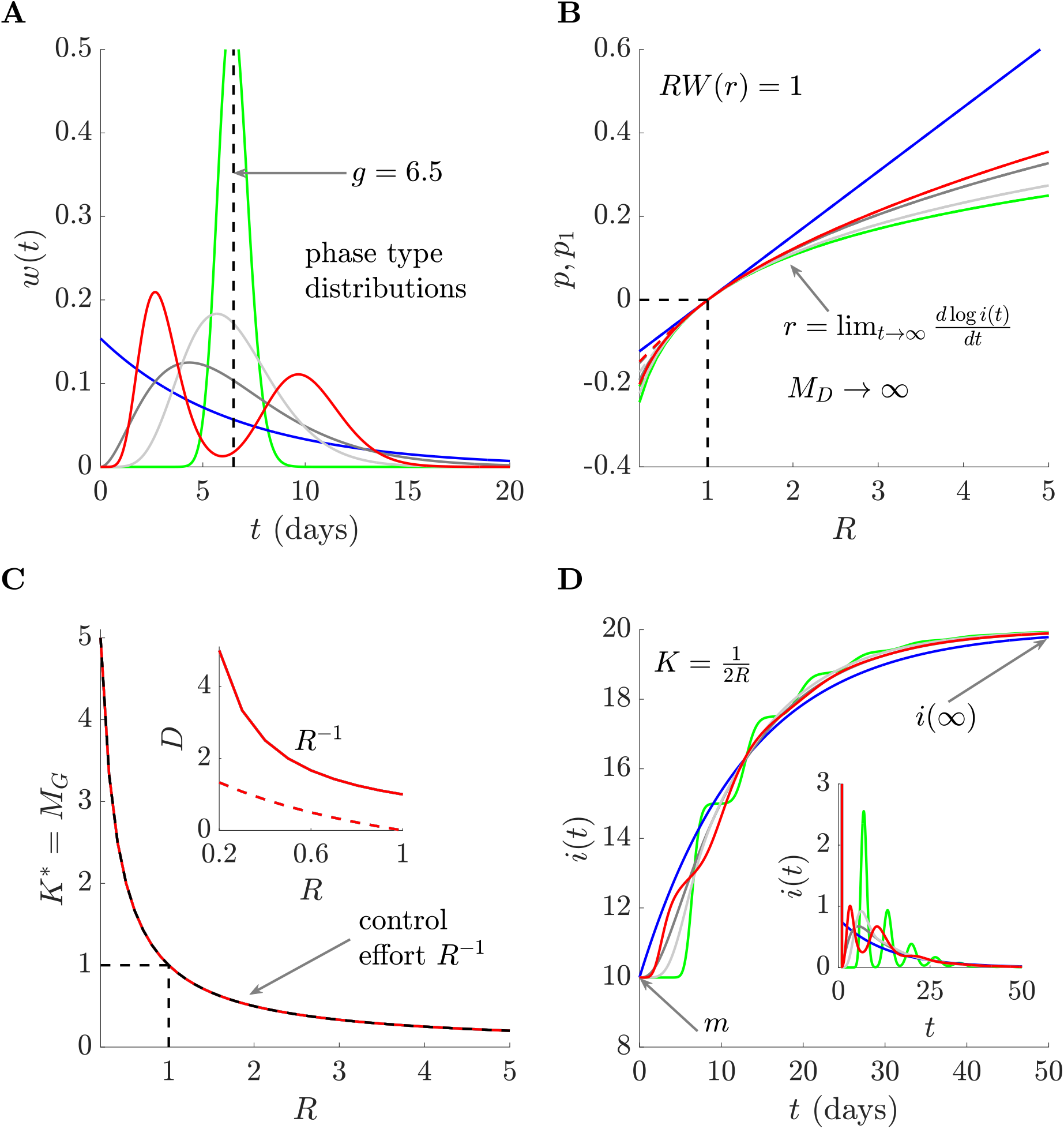
Epidemic controllability under ideal conditions. We assess controllability via gain and delay margins for epidemics subject to constant (non-dynamical) control *K*(*s*) = *k* with phase crossover frequencies of 0 (see text). Panel A shows the generation time distributions *w*(*t*) of simulated epidemics that we analyse, which have fixed mean generation time *g* (taken from COVID-19 [3]) but feature markedly different shapes. Panel B plots the growth rate *r* of these epidemics (colours match panel A), which is the dominant pole *p* (solid) of the resulting TFs *G*(*s*). These strongly match the dominant pole *p*_1_ (dashed) of an approximating epidemic described by 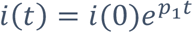. Panel C plots the gain margin *M*_*G*_ or critical controller *K*^*^ that drives the system to the brink of instability (the delay margin *M*_*D*_ here is infinite). The *K*^*^ curves from every *w*(*t*) exactly equal *R*^−1^. These curves correlate well with *D* (inset), the Euclidean distance between *L*(*s*) and −1. Panel D demonstrates that although controllability is the same, transient dynamics of infections may differ (they also depend on non-dominant system poles). We plot incident infections *i*(*t*) in response to stable numbers (main) of imported infections (*m*(*t*) = *m*) and to a 1-day pulse (inset) of *m* imports (colours match panel A).

The dominant pole and hence the effective growth rate also shifts, from being the solution of *RW*(*s*) = 1, to that of (*kR*)*W*(*s*) = 1. As this equation is only scaled, the growth rate is now related to the effective reproduction number *kR*. For gamma distribution generation times with parameters (*a, b*) for example (see Methods), the growth rate changes from 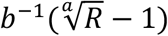 to 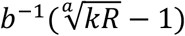. Consequently, if *ω*_PC_ = 0, we can completely describe the controllability of an epidemic using the size of reproduction numbers or growth rates. As growth rates are asymptotic (i.e., other poles decay in impact as *t* → ∞) we can equally describe controllability from exponential growth models that approximate complex renewal processes (**Fig 2B**). Note that epidemics with the same controllability may have diverse responses to imports (**Fig 2D**).

Our framework therefore supports the conventional definition that larger *R* or *r* indicates lower controllability but reveals that this requires *ω*_PC_ = 0 and that our controller is constant (i.e., we need *k*(*t*) = *kδ*(*t*)). Under these conditions we cannot destabilise the epidemic through perturbations that only add delay (or change phase). This holds for broad classes of phase-type generation time distributions. We show next that our more generalised controllability definitions are necessary because these settings are strongly restrictive and not likely to occur in practice i.e., control actions frequently introduce dynamics (e.g., by modifying incubation periods, generation times and infectiousness durations [21,22]). Further, we know that delays to interventions, surveillance biases, presymptomatic spread and superspreading, all impact controllability. We demonstrate that our definitions can rigorously unify these complexities.

### Problems with existing controllability definitions

Previously, we established conditions under which our generalised framework for assessing controllability reduces to the popular but informal definition applied in epidemiology. However, the conditions that allow this interpretation are strongly restrictive for two reasons. First, the only controller guaranteed to satisfy |*K*(0)| = *R*^−1^ and have unique *ω*_PC_ = 0 is the constant *K*(*s*) = *k*. This controller seems unrealistic given that interventions not only scale infections but also change the distribution of generation times and other epidemiological quantities and hence induce additional dynamics (and poles in *G*(*s*)) [20–22]. Any realistic intervention (e.g., social distancing or contact tracing) likely scales infections and slows them from occurring.

We demonstrate this for the generation time distributions in **Fig 2A** using controllers of form 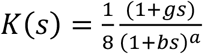, which induce minimal dynamics and satisfy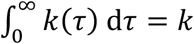. Here *K*(*s*) can model interventions that change the effective reproduction number as well as the generation times of the epidemic. For example, if the uncontrolled epidemic has exponentially distributed *w*(*t*) with mean *g* then 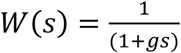 and the loop TF changes from 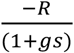 to 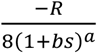 i.e., the control scaled down infections by a factor of 8 and forced the mean generation time to *ab*. This illustrates how controllers can realistically alter dynamics. Intervention-driven changes to generation times have been observed for malaria, COVID-19 and other diseases [19,20].

When *K*(*s*) is applied to epidemics with *R* = 4, if *ω*_PC_ = 0, then *M*_G_ = |−*K*(0)*R*|^−1^ = 2. This controller is strongly stabilising (we can double infections before critical stability), attenuating infections so that the effective reproduction number of the controlled epidemic is 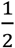. However, this standard interpretation is misleading and incomplete due to the temporal variations that control actions may introduce. In **Fig 3A** we compare the resulting epidemic trajectories under two *K*(*s*) examples. For the first (*b* = 2, *a* = 4) the gain and delay margins together with the response to a stable input of *m*(*t*) = 100 infections over time is consistent with **Fig 2**. Note that the *g* and *R* we use here are among values estimated for COVID-19.

**Fig 3:**
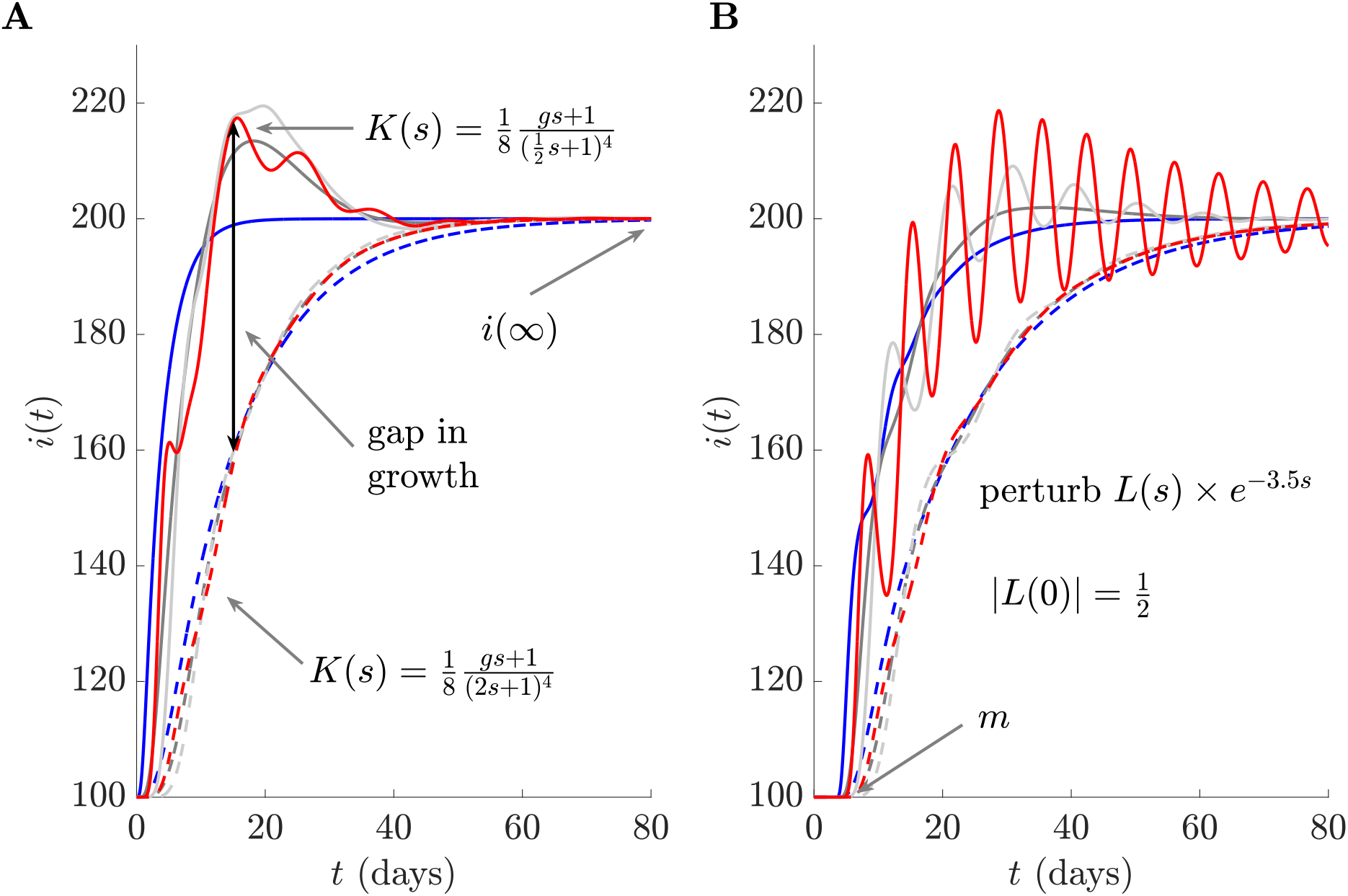
Controllers introducing additional dynamics. We simulate epidemics that are forced by a constant supply of imported infections (*m*(*t*) = 100). Panel A shows the resulting curves of incidence for epidemics with generation time distributions from **Fig 2** (excluding the green one because this becomes unstable, curves match in colour) when non-constant control *K*(*s*) is applied. There are major discrepancies among responses to interventions (controllers) that induce substantial dynamics (solid) and those behaving as we would conventionally expect (dashed). The former show salient transients that disrupt controllability and feature finite delay *M*_*D*_ (and in some cases *M*_*G*_ < 2). Conventional interpretations expect *M*_*G*_ = 2, *M*_*D*_ → ∞. The long-term incidence *i*(∞) remains, however, unchanged for all our controllers. In panel B we apply a 3.5-day delay (*e*^−3.5*s*^ in the *s* domain). This pushes the curve from panel A with finite *M*_*D*_ ≈ 4.2 days towards instability. The value of a two-margin description is clear.

Strikingly, for the second case 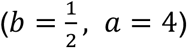, the response is markedly different, featuring oscillations and faster (transient) growth that might initially strain available resources. The gain margin for these cases is still 2 (though in some instances it can fall below 2) but, importantly, the delay margin for one of the *w*(*t*) in **Fig 2A** becomes finite and small (*M*_D_ ≈ 4.2 days). This effect is pronounced and this *K*(*s*) can cause the overall generation time to shrink to roughly 2 days. Case isolation was found to cause similar shrinking for COVID-19 [21]. Smaller *M*_D_ values can occur for further *w*(*t*) types under more complex controllers (not shown). As much is unknown about these rebound effects of interventions [22], we cannot be certain about realistic formulae for *K*(*s*). Recent works [35] emphasise the need for collecting the data types that will allow precise *K*(*s*) parametrisation. When such data become available, our margins will be best placed to assess controllability and expose any unexpected rebound effects.

The finite delay margin is especially valuable, revealing that interactions between the epidemic and interventions can cause robustness losses. Real interventions always have latencies [13], making *M*_D_ crucial. If control is applied after a 3.5-day delay, we obtain infection curves as in **Fig 3B**. There we observe that the red curve approaches instability and realise that there is a hard limit from *M*_D_ on how late we can respond to an epidemic if we want control to work. The importance of delays in epidemic control is a known issue [9,16] but it is rarely factored into epidemic controllability directly. Our (*M*_D_, *M*_G_) framework is comprehensive and exposes the pitfalls of measuring controllability only in terms of *R* or *r* (while not shown, the dominant poles and hence *r* in **Fig 3** are similar for both the finite and infinite *M*_D_ scenarios).

The second major problem with conventional definitions of controllability is that they are not easily computed, interpreted or compared when practicalities such as presymptomatic spread, superspreading, variant dynamics and surveillance imperfections (e.g., reporting delays and incomplete case ascertainment) occur [1,24]. In the next two sections, we expand our models and demonstrate that the (*M*_D_, *M*_G_) framework presents a unified and interpretable approach to measuring and monitoring epidemic controllability under all of these complexities. No matter the specific model structure, the boundaries of controllability specified by our (*M*_D_, *M*_G_) pair are directly comparable and possess exactly the same interpretation as in **Fig *1***.

### Surveillance limitations and presymptomatic spread

Until now we have assumed that we can observe and apply control to all new infections. This is unrealistic as commonly we can only count cases or deaths, which are delayed and scaled versions of infections [36,37]. Here we generalise **Eq. (1)** and **Eq. (2)** to include these effects. We denote the proportion of infections that we observe as cases by probability 0 ≤ *ρ* ≤ 1 and model the latency in observing these cases with a distribution *h*(*t*). Our controller acts on the incidence of cases *c*(*t*), and *i*(*t*) − *c*(*t*) infections remain unobserved. This yields **Eq. (3)**.

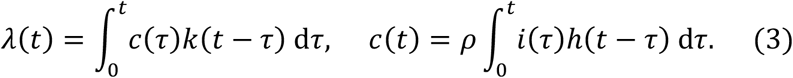

The unobserved infections continue to propagate the epidemic as they remain uncontrolled. We therefore construct the combined renewal model of **Eq. (4)** below.

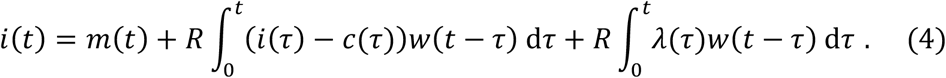

This collapses into **Eq. (1)** when reporting is perfect i.e., *ρ* = 1 and *h*(*t*) has all its probability mass at the present (*h*(*t*) = *δ*(*t*), the Dirac delta) so that *c*(*t*) = *i*(*t*).

We again take Laplace transforms of **Eq. (3)** and **Eq. (4)** to obtain our key TFs for evaluating epidemic controllability in **Eq. (5)**. We illustrate this architecture in **Fig 4A** and observe that we also obtain TFs for the observed cases easily since *C*(*s*)*M*(*s*) ^−1^ = *ρH*(*s*)*G*(*s*).

**Fig 4:**
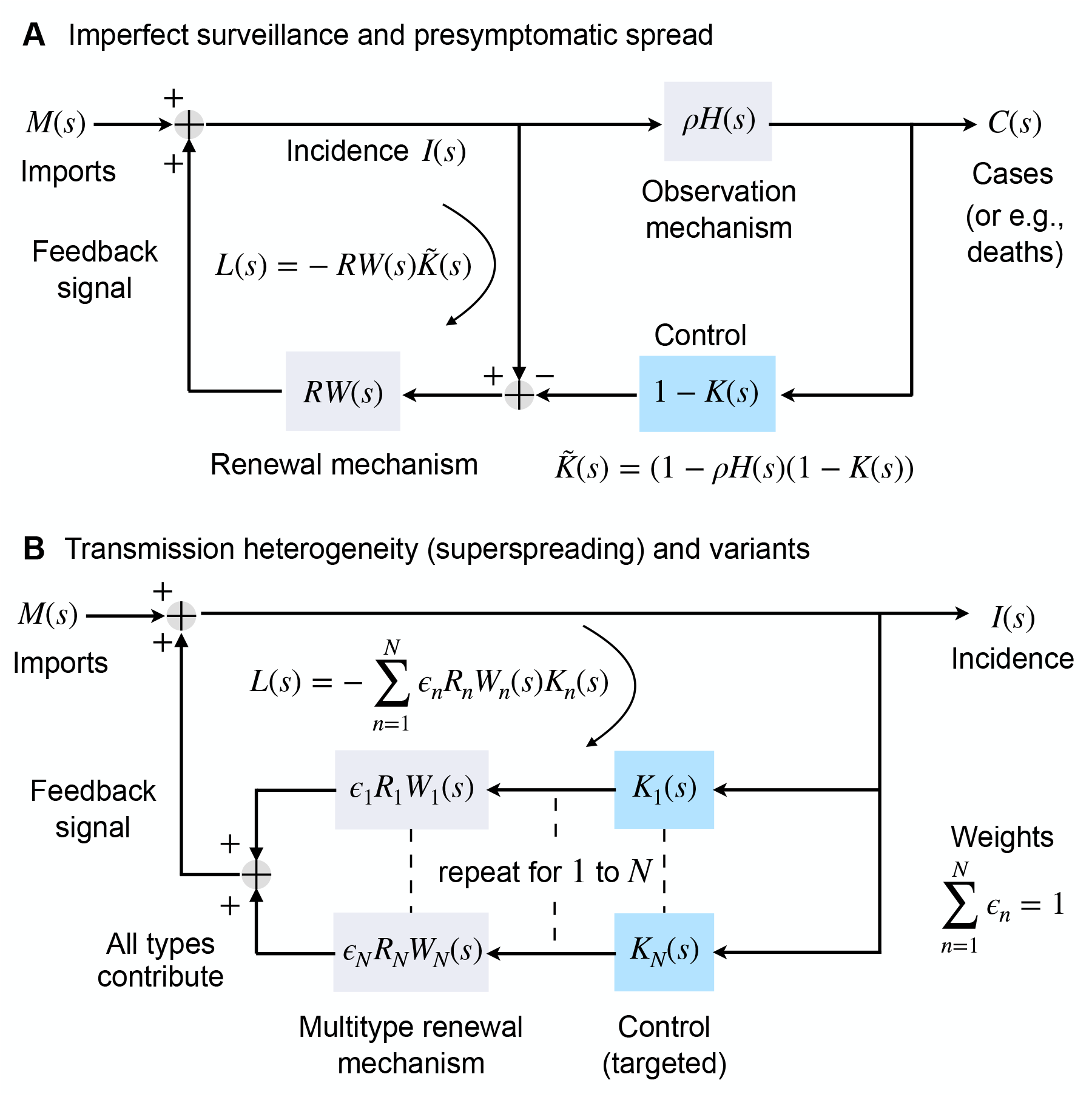
Generalised controlled renewal model architectures. Panel A illustrates the block diagram of a renewal model for which only a portion of the new infections *I*(*s*) are observable and hence can be controlled by *K*(*s*). This portion *C*(*s*) may model cases, deaths or any other time series that is mediated by a scale factor *ρ* and a lag distribution *H*(*s*). This architecture represents imperfect surveillance mechanisms or presymptomatic spread. Panel B shows the structure of a multitype, controlled renewal model describing *N* infectious types or stages with diverse reproduction numbers *R*_*n*_ and generation time distributions *W*_*n*_(*s*). The weight *ϵ*_*n*_ is the fraction of new infections of type *n*. This architecture models transmission heterogeneity including superspreading, co-circulating variants and diseases with multiple routes for spread but considers the combined contributions of all types (hence contact matrices are not needed). Both panels have closed loop TFs *G*(*s*) = *I*(*s*)*M*(*s*) ^−1^ = X1 + *L*(*s*)Y^−1^, with loop TF *L*(*s*) as described. See main text for details on how *K*(*s*) and the *K*_*n*_(*s*) define controllability.

**Fig 5:**
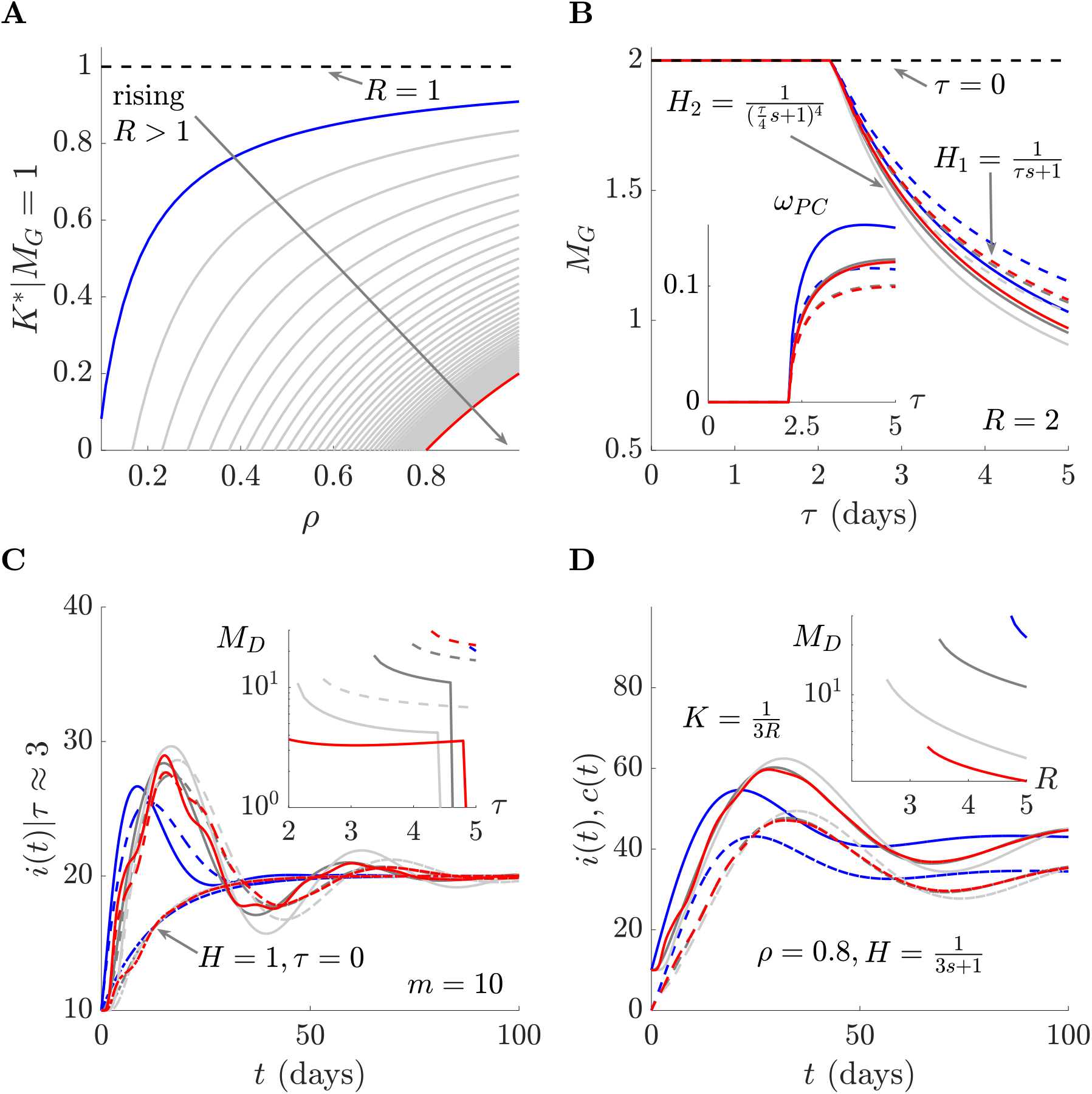
Surveillance noise and presymptomatic spread. We investigate how imperfect case reporting, or equivalently presymptomatic spread, limits the controllability of epidemics using our (*M*_*D*_, *M*_*G*_) framework. Panel A shows for curves of constant *R* ≥ 1 (rising from blue to red, which is at *R* = 5) how the reporting rate or proportion of symptomatic infections, *ρ*, reduces controllability. Smaller *ρ* requires more control effort to attain critical stability i.e., a smaller *K*^*^ is needed for a gain margin *M*_*G*_ = 1. There is no reporting delay or presymptomatic distribution in this analysis so *H*(*s*) = 1. Panel B sets *ρ* = 1 and investigates the influence of two *H*(*s*) distributions, *H*_1_ (dashed) and *H*_2_ (solid) modelling exponential and gamma distributions. Both have mean lag *τ, R* = 2 and a controller applied that achieves *M*_*G*_ = 2, if the phase crossover frequency *ω*_PC_ = 0. We find that as *τ* increases *M*_*G*_ < 2 indicating a decline in controllability. This results from *ω*_PC_ increasing above 0 (inset). Colours in this and panels C-D match the generation times modelled from **Fig 2A** (excluding the green). Panel C confirms *H*(*s*) causes the delay margin *M*_*D*_ to become finite (inset, dashed or solid corresponding to panel B). This reduced controllability is visible from the peaked, oscillatory response in new infections *i*(*t*) for a constant number of imports *m*(*t*) (main). This effect is similar to that in **Fig 3**. Here dot-dashed lines plot the response in the absence of *H*(*s*). Panel D shows the combined influence of lags and under-reporting given the constant controller of 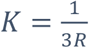. The inset demonstrates how *M*_*D*_ falls with *R* and the main shows the infection (solid) and case *c*(*t*) (dashed) epidemic curves in response to constant imports (colours match generation time distributions).

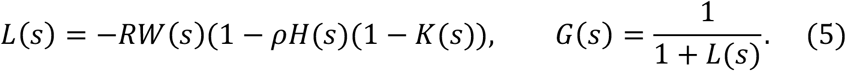

When *K*(*s*) = 1 in **Eq. (5)** we recover the uncontrolled epidemic TFs (see **Eq. (M1)**). Perfect surveillance means *ρH*(*s*) = 1 and reverts **Eq. (5)** to **Eq. (2)**. If we instead perform control on another proxy of infections, for example deaths or hospitalisations, then *ρ* is the proportion of infections that lead to mortality or hospitalisation (e.g., for the incidence of deaths this includes the infection fatality ratio and the proportion of deaths that are observed). The distribution *h*(*t*) then models the lag from becoming infected to mortality or being admitted to hospital [37,38].

This formulation equally models presymptomatic and asymptomatic spread, with *h*(*t*) defining the delay between infection and presenting symptoms and *ρ* as the proportion of infections that never become symptomatic. We compute our (*M*_D_, *M*_G_) pair to assess how these differing transmission and surveillance characteristics impact controllability. **Eq. (5)** includes all the key controllability factors outlined in [1] and describes targeted interventions such as quarantine, contact tracing or isolation but not widescale lockdowns (we only control observed infections). Lockdowns and other non-selective interventions conform more closely to **Eq. (2)** as they act indiscriminately on all infections, including those that we never observe.

We know from earlier that critical stability is achieved when *L*(*s*) = −1. We substitute this into **Eq. (5)** and find that our control needs to satisfy the left side of **Eq. (6)**. As a constant *K*(*s*) = 0 represents the maximum possible control effort (i.e., all observed infections are suppressed completely), we insert this condition and rearrange to derive the threshold on the right side of **Eq. (6)**, outlining the requirements on the surveillance noise or level of presymptomatic spread for the epidemic to just be controllable. A smaller |*ρH*(*s*)| causes loss of controllability and provides evidence that widescale interventions or surveillance improvements are needed. The relations of **Eq. (6)** are only required to hold at the *s* = *jω* satisfying *L*(*s*) = −1.

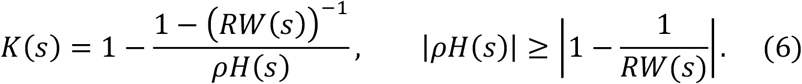

If *ω*_PC_ = 0 then this requirement is met at *ρ* ≥ 1 − *R*^−1^, as *W*(0) = *H*(0) = 1. This matches the critical contact tracing efficiency derived in [2] and the presymptomatic condition of [1] and confirms how our methodology generalises more conventional notions of controllability (it also relates to the herd immunity threshold though we do not consider this directly as our models neglect depletion of susceptibles). **Eq. (6)** verifies that we need both margins because *ω*_PC_ = 0 is not guaranteed here, even if controllers are constant. The temporal impact of imperfect surveillance or presymptomatic spread via *H*(*s*) means that the dynamics leading to situations as in **Fig 3** always exist. Transient dynamics are crucial and unavoidable.

We verify this point in **Fig 5**, showing how controllability depends on *ρ* and *H*(*s*). We first set *H*(*s*) = 1 and explore the controller gain needed to get *M*_*G*_ = 1, which sets critical stability. In the absence of under-reporting, we have *ρ* = 1 and *K*^*^ = *R* ^−1^ for any *R*. **Fig 5A** shows that our required *K*^*^ substantially deteriorates, highlighting that we need additional control effort to stabilise the epidemic as *ρ* decreases. When *K*^*^ = 0, the epidemic is no longer controllable by these targeted interventions. If we cannot improve surveillance quality or, equally diminish asymptomatic spread (so *ρ* rises), then population-level controls are warranted. Strikingly, at *R* = 5 (red), we cannot control the epidemic unless more than 80% of all new infections are observed (sampled) or symptomatic. **Eq. (6)** defines fundamental limits on controllability.

In **Fig 5B** and **Fig 5C** we assume perfect reporting and test the influence of delays in reporting or equivalently lags in infections becoming symptomatic. We investigate two *h*(*t*) distributions, *H*_1_(*s*) and *H*_2_(*s*) in the frequency domain, with results respectively as dashed or solid. These, model exponential and gamma distributed delays with means *τ*. We apply controls that force *M*_*G*_ = 2 when *ω*_PC_ = 0 but find in **Fig 5B** that our gain margin declines with *τ*. This occurs as *ω*_PC_ > 0 (inset). **Fig 5C** further shows that the delay margin *M*_*D*_ becomes finite, decaying with *τ* (inset). Hence, *H*(*s*) reduces both the scaling and delays that the controlled epidemic can robustly support. Incident infections *i*(*t*) display oscillatory dynamics with substantial peaks (main). This contrasts the plots featuring no delay i.e., *τ* = 0 (dot-dashed). Colours indicate the *w*(*t*) from **Fig 2A** underlying results in **Fig 5B, Fig 5C** and **Fig 5D**.

On its own, *H*(*s*) substantially reduces our controllability. At *τ* ≥ 4 we find that *M*_D_ → 0 (and *M*_G_ < 1) signifying that the epidemic is now unstable. Epidemics with larger *τ* are necessarily uncontrollable. We combine both *ρ* and *H*(*s*) in **Fig 5D** but vary *R* and apply a strong controller that scales down cases by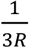. Even for this constant control we find a finite *M*_D_ that declines with *R* (inset) and large amplitude oscillations in *i*(*t*) (solid, main). We also plot the observed cases *c*(*t*) (dashed), which are the fraction of infections we can control. Both of the (*M*_D_, *M*_G_) pair are therefore critical to accurately quantifying epidemic controllability. In **Fig 5** the ranges of *ρ* and *τ* that we explore are realistic and even better than those often reported for countries with good COVID-19 surveillance, which can feature smaller *ρ* or larger *τ* [37,39,40].

### Superspreading, variants and multiple infector types

Our (*M*_D_, *M*_G_) framework can also evaluate the controllability of epidemics that are composed of multiple infectious types or transmission routes. This models superspreading, co-circulating variants and pathogens with multiple pathways of spread. We unify these multitype epidemics using the renewal process of **Eq. (7)**, which features *N* distinct types or pathways.

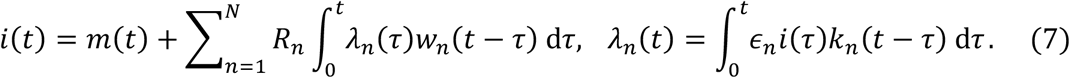

We denote the reproduction number, generation time distribution and controller of the *n*^th^ type with subscript *n*. The parameters *ϵ*_*n*_ define the proportion of incidence associated with the *n*^th^ type and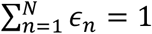. By dividing control into *N* functions, we allow for type-specific control.

This includes non-targeted control (all *k*_*n*_(*τ*) are the same) and situations where some types are uncontrolled (those *k*_*n*_(*τ*) = 1), perhaps due to being unobservable.

Specialisations of **Eq. (7)** can model superspreading or transmission heterogeneity (e.g., we set 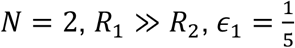 and 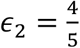 to describe cases where 20% of new infections have substantially larger transmissibility, which aligns with data on many diseases [41]), pathogenic variants with differing transmissibility and generation times (e.g., with *N* as the number of co-circulating variants, although we assume early growth so that the *ϵ*_*n*_ are fixed [42,43]) and diseases with diverse transmission pathways (e.g., Ebola virus disease has sexual and non-sexual pathways with distinct *w*_*n*_(*t*) [30]). These models do not include explicit interaction among types (though all types compose *i*(*t*)) as this commonly requires additional cross-type reproduction numbers and auxiliary data (e.g., to construct contact matrices or to calculate epidemic thresholds) [44,45]. Such extensions are possible provided the interacting system can be framed as a multidimensional, linear renewal model. While we do not consider these extensions, if appropriately formulated our margins should remain valid.

We take Laplace transforms of **Eq. (7)** to construct **Eq. (8)**, which is amenable to our gain and delay margin controllability analyses. We sketch the architecture of this model in **Fig 4B**.

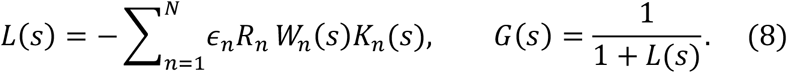

Using the fact that *W*_*n*_(0) = 1, we find that if *ω*_PC_ = 0 then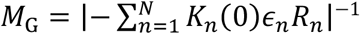. We can therefore scale the epidemic by a quantity that is a weighted sum of control, reproduction numbers and proportions of the contributing infectious types. As we showed in above sections, this condition is only likely to be met if every controller is constant (at which also *M*_D_ → ∞). If controllers introduce dynamics, which is realistic, then we expect effects similar to **Fig 3**.

**Eq. (8)** provides the flexibility to investigate several controllability problems. We focus on two questions about the limitations of targeted control for heterogeneous populations. We let *N* = 2 and assume that *R*_1_ ≥ *R*_2_ so that type 1 represents individuals with the more transmissible variant or superspreading nodes. We consider non-selective control where *K*_1_(*s*) = *K*_2_(*s*) = *K*(*s*) and targeted control, in which only one type is controlled. We only target type 1, which is more transmissible, so type 2 is uncontrolled and *K*_2_(*s*) = 1. Our first question asks under what conditions the targeted approach, which is often proposed as an efficient control scheme [11,41], fails to suppress the overall epidemic, making non-selective control unavoidable.

For this two-type epidemic *L*(*s*) = −(*ϵ*_1_*R*_1_*K*_1_(*s*)*W*_1_(*s*) + *ϵ*_2_*R*_2_*W*_2_(*s*)) for targeted control and −*K*(*s*)(*ϵ*_1_*R*_1_*W*_1_(*s*) + *ϵ*_2_*R*_2_*W*_2_(*s*)) for non-selective control, with *ϵ*_2_ = 1 − *ϵ*_1_. In both cases, *ω*_PC_ = 0 and *W*_1_(0) = *W*_2_(0) = 1 (see Methods). If we only apply constant controllers, then *M*_D_ → ∞ and controllability is exclusively defined by the values of *M*_G_, which are computed as |*ϵ*_1_*R*_1_*K*_1_(0) + *ϵ*_2_*R*_2_|^−1^ and |*K*(0)|^−1^|*ϵ*_1_*R*_1_ + *ϵ*_2_*R*_2_|^−1^. To attain some specific *M*_*G*_, we require 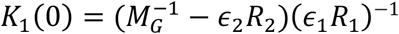 and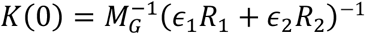. We can combine these relations to get the left side of **Eq. (9)**, which shows how much smaller *K*_1_(0) needs to be than *K*(0) i.e., how much more targeted control effort is required to attain our desired *M*_G_.

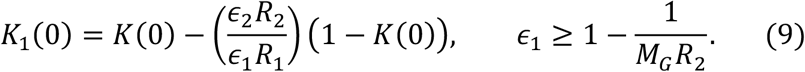

We plot the control efforts *K*^*^ = *K*(0) and 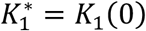 from both strategies that are necessary to achieve critical stability (*M*_G_ = 1) in **Fig 6A**. There we observe the limits of targeted control as a critical *ϵ*_1_ cut-off (dashed vertical). This follows from the positivity constraint 0 ≤ *K*_1_(0) < 1, where 1 is no control and 0 defines perfect control, in which type 2 infections are neutralised. We derive this for any desired gain margin on the right side of **Eq. (9)**. Interestingly, this cut-off does not depend on *R*_1_ and, if *M*_G_ = 1, it indicates that targeted control only works when the proportion of superspreading nodes or type 1 variants is above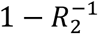. This procedure is easily generalised to *N*-type epidemics where we can control a subset *χ* of the types. The controllability cut-off then requires the uncontrolled proportion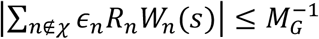.

**Fig 6:**
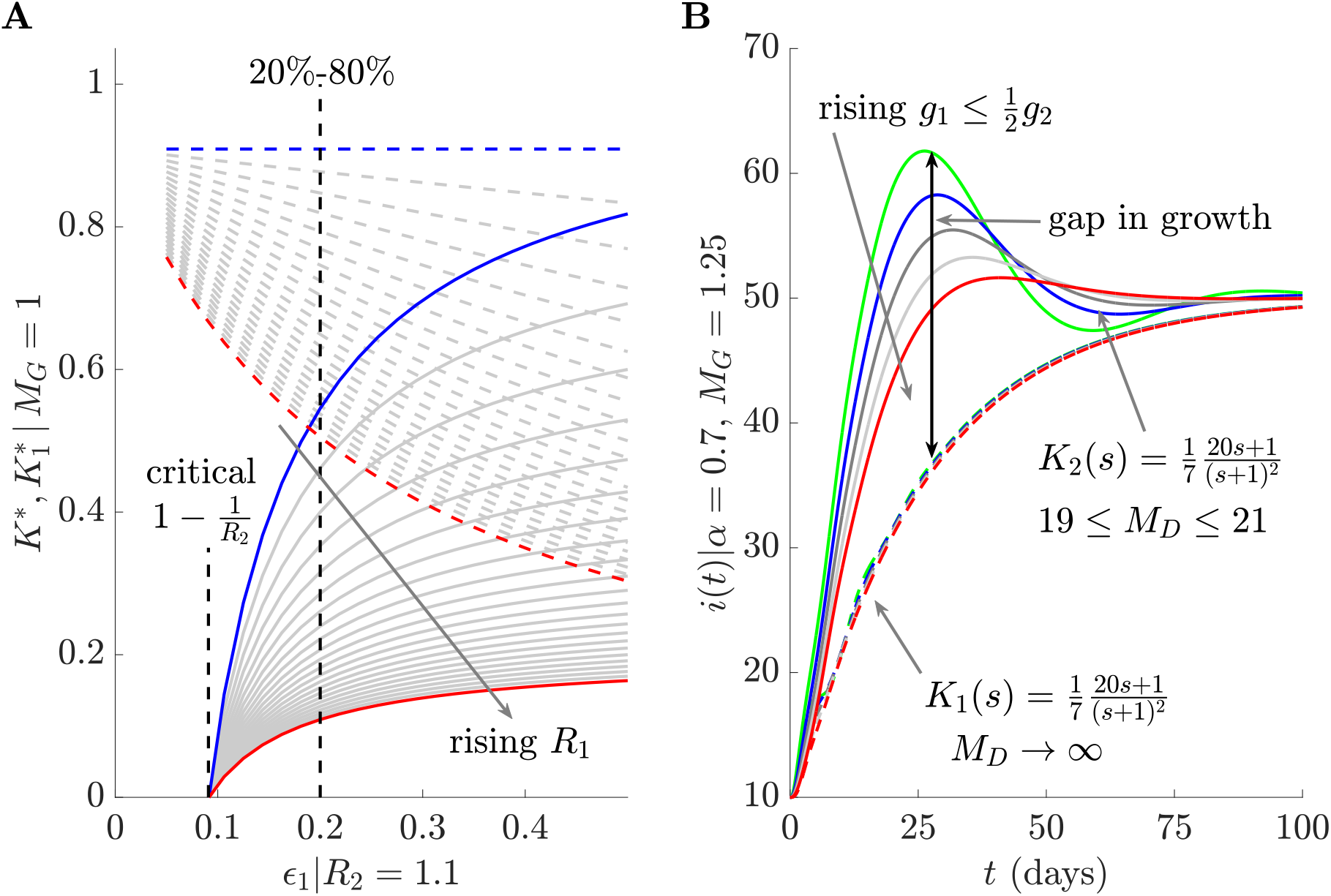
Targeted control in multitype epidemics. We explore controllability and performance limits for epidemics that involve two distinct types, modelling superspreading or co-circulating variants. Panel A plots the constant control effort necessary for critical stability (*M*_*G*_ = 1) under a non-selective strategy with controller *K*^*^ that reduces infections of both types (dashed) and a targeted strategy with controller 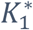 that only reduces infections of type 1 (solid), which has larger transmissibility *R*_1_ ≥ *R*_2_ = 1.1. For both strategies, we vary the proportion of type 1, *ϵ*_1_, and curves are for increasing *R*_1_ from blue (1.1) to red (5.5) with intermediate values in grey. We use a vertical line to show the *ϵ*_1_ for the commonly used 20-80 superspreading rule that describes realistic epidemic heterogeneity. Targeted control requires substantially more effort (as it must also account for the uncontrolled type 2), and the epidemic is uncontrollable if *ϵ*_1_ is smaller than the critical vertical line (see **Eq. (9)**). Panel B considers targeted controllers that introduce dynamics and only apply *K*_1_(*s*) or *K*_2_(*s*) to reduce either type 1 or 2 infections. We fix *ϵ*_1_*R*_1_ = *ϵ*_2_*R*_2_ = *α* so that all types contribute to overall transmissibility equally and both controllers lead to the same *M*_*G*_ > 1. We illustrate how new infections *i*(*t*) change due to both schemes (dashed and solid respectively), where type 1 is the faster variant possessing mean generation time *g*_1_ ≤ *g*_2_ = 8 days. Targeting the slower type 2 leads to worse performance (including faster transient growth) and is sensitive to *g*_1_ (curves are not grouped).

Our second question relates to the interaction between differing generation times of the types and induced controller dynamics. We consider targeted control of either type or variant with type 1 having smaller mean generation time and hence being faster than type 2 i.e., *g*_1_ ≤ *g*_2_. We set *ϵ*_1_*R*_1_ = *ϵ*_2_*R*_2_ = *α* to remove any relative transmissibility advantage between the types. Consequently, variations in the infections caused by the types emerge from their generation time distribution differences. Targeted control applies non-constant control *K*_1_(*s*) exclusively to type 1 or *K*_2_(*s*) exclusively to type 2, yielding loop TFs *L*(*s*) = −*α*(*K*_1_(*s*)*W*_1_(*s*) + *W*_2_(*s*)) and −*α*(*W*_1_(*s*) + *K*_2_(*s*)*W*_2_(*s*)). Because the controller induces additional dynamics, we are neither guaranteed *ω*_PC_ = 0 nor *M*_D_ → ∞ and must evaluate the complete (*M*_D_, *M*_G_) pair.

We compute these margins and dynamical responses to constant importations in **Fig 6B** for a range of fast type 1 generation time distributions *w*_1_(*t*) and a fixed (slow) type 2 distribution *w*_2_(*t*). Although *M*_G_ is the same for both schemes, controlling type 2, which may occur when transmission chains of slower variants are easier to interrupt, yields worse performance. The overshoots and oscillations are also accompanied by a finite *M*_D_, highlighting that neglecting the faster variant can potentially reduce robustness of the controlled epidemic to perturbations or equally reduce controllability below what we may expect from conventional measures based on reproduction numbers or asymptotic growth rates. For certain controllers (not shown) we also find that *ω*_PC_ > 0 can occur and reduce *M*_G_ for either targeted scheme. This underscores the importance of our two-margin solution to understanding controllability.

## Discussion

Measuring the controllability of an infectious disease subject to various intervention options is a fundamental contribution of mathematical modelling to epidemiology [4,12]. However, there exists no rigorous and precise definition of what controllability means [8,10] and studies have highlighted a need for robust analytical frameworks to better appraise the impacts of targeted and reactive interventions [1]. Currently, the distance from the epidemic threshold of *R*=1 or *r*=0 is frequently used to measure controllability. Here we have demonstrated that this notion of controllability, although reasonable, is idealistic and likely misleading because neither *R* nor *r* completely and unambiguously measures distance from stability. We proposed an alternative and analytic definition of this distance by reformulating the disease transmission process as a positive feedback loop and leveraging results from control engineering [31].

We derived epidemic transfer functions to describe the dynamics of this loop and model how stabilising interventions interrupt and attenuate this positive feedback (e.g., by blocking new infections through quarantines). For already stable or controlled epidemics, we test robustness to perturbations or uncertainties that amplify infections along this loop (e.g., by relaxing any interventions or from pathogenic variants). This allowed us to develop stability margins that accurately measure the distance from stability (**Fig *1***) in units of the scale and speed of the required control efforts. The gain and delay margins are key metrics from control engineering [17,18], a field that studies stability and feedback problems across many dynamical systems. Although there is increasing interest in using tools from this field to better understand infectious disease spread [9,46–49], our study appears to be among the earliest to construct margins for epidemics and appraise existing notions of disease controllability.

Our central contribution is a flexible method for quantifying epidemic controllability that is both computable and easily interpreted across many salient characteristics of infectious diseases. This is important for three main reasons. First, *R* and *r* can lose their meaning or comparability as threshold parameters when characteristics such as superspreading and multitype spread are included [24,50]. Second, for a given transmission model there can be numerous ways of constructing and defining valid epidemic thresholds and these are not always consistent when assessing interventions [25,45,50]. For example, when interventions change generation times then we can find situations where *r* increases yet *R* decreases [51]. Third, earlier frameworks were unable to directly include reactive or feedback effects within their measures and did not account for how the implementation of interventions might modify effectiveness.

In contrast, our gain and delay margins maintain their interpretation, validity, uniqueness and comparability across complex disease models and explicitly reflect feedback loops intrinsic to transmission and intervention. These properties allowed controllability to be measured across realistic generation time distributions (**Fig *2***), constraints on interventions (**Fig *3***), surveillance imperfections (**Fig *5***) and transmission heterogeneities (**Fig 6**). Principal insights emerging from this unified approach were that (i) *R* and *r* only track controllability in restrictive settings where interventions do not alter temporal disease characteristics and are applied instantly, (ii) sharp thresholds of controllability exist due to presymptomatic spread, superspreading, delays and under-reporting and co-circulating variants that generalise 1-1/*R* type results and (iii) the delay margin is crucial because lags along feedback loops (from both intervention delays and surveillance biases) can destabilise epidemics that are conventionally deemed controlled.

While our approach rigorously incorporates many realistic epidemic complexities and extends earlier frameworks [1,8,10], it depends on several simplifying assumptions, which we made to ensure tractability and to extract general insights. Specifically, our analysis uses deterministic renewal models and assumes constant *R* or *r*. Although some or all of these assumptions are common to seminal studies and recent works on controllability thresholds [1,43], the influence of stochasticity in disease transmission can be substantial [7,11]. We recommend computing our margins to initially assess intervention impact and then using them to guide the running of more complex stochastic models. Our margins are only well-defined for linear systems, which include epidemics describable by renewal models with constant *R*. If *R* varies on the timescale of interventions or involves non-linear effects such as saturation or susceptible depletion, this assumption may be invalidated. However, we can use piecewise-constant transmissibility approximations and fit renewal models to each piece, to partially circumvent this issue.

Moreover, we examine linear and reactive control actions only (i.e., convolutions of kernels with past infections). This improves upon many studies, where controllers simply multiply and reduce *R* or *r* but may not model other notable types of interventions, such as those reducing infections due to non-linear switching triggers or those that completely ignore feedback signals in favour of predetermined action [34,52]. Understanding the relative benefits of these different strategies is an ongoing area of research. Last, we comment that controllability here focussed on intrinsic epidemic dynamics and neglected the costs of actions. Including how these costs further constrain the realisable limits of controllability, as well as incorporating key behavioural effects within our feedback loops are the future directions of this research.

In summary, we demonstrate that controllability is only completely and accurately measured by the distance of the loop transfer function *L*(*s*) from −1. This generalises and improves upon the conventionally used distances of *R* from 1 or *r* from 0, but still admits interpretable margins or safety factors that quantify how much we can scale infections or delay interventions to attain critical stability. This allows us to better evaluate when targeted interventions are insufficient and hence when non-selective controls such as lockdowns are justified from the viewpoint of curbing transmission. We find that targeted controls fail when the dynamics of the unobserved or untargeted infectious population, together with constraints on surveillance and intervention implementation cross margin thresholds that are analytically derived from our framework.

## Data Availability

This study produced no data. All code underlying the analyses and figures within this paper will be made freely available in MATLAB at: https://github.com/kpzoo/EpidemicControllability

## Funding

KVP acknowledges funding from the MRC Centre for Global Infectious Disease Analysis (reference MR/X020258/1), funded by the UK Medical Research Council (MRC). This UK funded award is carried out in the frame of the Global Health EDCTP3 Joint Undertaking. The funders had no role in study design, data collection and analysis, decision to publish, or manuscript preparation. For the purpose of open access, the author has applied a ‘Creative Commons Attribution’ (CC BY) licence to any Author Accepted Manuscript version arising from this submission.

## Data Availability

All data and code underlying the analyses and figures within this study are freely available in MATLAB at: https://github.com/kpzoo/EpidemicControllability

## Author Contributions

**KVP** – Conceptualisation, Formal Analysis, Funding Acquisition, Investigation, Methodology, Project Administration, Software, Validation, Visualisation, Writing – original draft, Writing – review and editing.

## Notes

### Competing Interest Statement

The authors have declared no competing interest.

### Funding Statement

KVP acknowledges funding from the MRC Centre for Global Infectious Disease Analysis (reference MR/R015600/1), jointly funded by the UK Medical Research Council (MRC) and the UK Foreign, Commonwealth & Development Office (FCDO), under the MRC/FCDO
Concordat agreement and is also part of the EDCTP2 programme supported by the European Union. The funders had no role in study design, data collection and analysis, decision to publish, or manuscript preparation. For the purpose of open access, the author has applied a Creative Commons Attribution (CC BY) licence to any Author Accepted Manuscript version arising from this submission.

### Summary of Updates

New analyses and figures. Text has also been revised to improve clarity.

